# Dose-response relationship of extended-release methylphenidate for ADHD in adults: post-hoc analysis based on a systematic review

**DOI:** 10.1101/2021.12.08.21267468

**Authors:** K Boesen, KJ Jørgensen, PC Gøtzsche

**Affiliations:** Meta-Research Innovation Center Berlin (METRIC-B); Cochrane Denmark; Institute for Scientific Freedom

## Abstract

**Objective:** To assess potential dose-response relationships of extended-release methylphenidate for ADHD in adults on efficacy outcomes.

**Study design and setting:** Post-hoc analysis based on a systematic review of extended-release methylphenidate (https://doi.org/10.1002/14651858.CD012857). Using data from clinical trials comparing multiple fixed-dose methylphenidate groups with placebo, we conducted subgroup meta-analyses for available efficacy outcomes.

**Results and conclusion:** Five trials used a fixed-dose design with multiple methylphenidate groups receiving different doses. All trials were pivotal industry sponsored studies conducted to obtain marketing authorisation. We analysed four efficacy outcomes: Self-rated ADHD symptoms (5 trials, 1807 participants), investigator-rated ADHD symptoms (5 trials, 1904 participants), quality of life (4 trials, 1158 participants), and peer-rated ADHD symptoms (2 trials, 879 participants). There were no dose-response relationships for any outcome.

## Introduction

In a recent letter^1^ commenting on our study^2^ of missing clinical trials in new drug applications of extended-release methylphenidate for attention deficit hyperactivity disorder (ADHD) in adults, the author wrote that, “there is substantial (if uncertain) body of evidence for the linear dose-response relationship” and that, “it is plausible that the doses used in the clinical trials have been too low.” The author of the letter referred to a systematic review^3^ and network meta-analysis of ADHD medications to support the statement. We did not find that the systematic review^3^ provided support to this idea. We therefore tested the dose-response hypothesis on a dataset we had recently collected for a systematic review on extended-release methylphenidate for adult ADHD.^4^

## Method

### Dataset

We identified clinical trials using a fixed-dose design including two or more methylphenidate groups and a placebo group. We collated data from all available efficacy outcomes reported separately for the methylphenidate groups. We did not include flexible dose titration trials, i.e. where participants are titrated to the highest tolerated dose, as this design is not appropriate to assess dose-response relationships. Data had been extracted already for our systematic review.^4^ For Weiss 2014, we included additional data from a Clinical Study Report^5^ published by Health Canada. The full dataset for this post-hoc analysis is available here: https://osf.io/wrf65/?view_only=ef3340ea339c44bdad5f83732cf4ba84.

### Analyses

We made meta-analyses in RevMan 5.4 using the fixed-effects model with inverse variance weighting. To avoid double counting of the placebo groups, we split the placebo comparator by keeping the mean value and variation and dividing the total number of placebo participants with the number of methylphenidate groups. Results were reported as mean differences. We reported the rating scale ranges in the plots.

## Results

We included five fixed-dose trials^6-10^ using four different extended-release formulations, each including between two and four fixed-dose methylphenidate groups (**Table 1**).

**Table 1.** Fixed-dose extended-release methylphenidate trials

| **Trial** | **Type of methylphenidate** | **Design** | **Groups** | **Reported outcomes** |
| --- | --- | --- | --- | --- |
| Spencer 2003<sup>6</sup> | Dex-methylphenidate extended-release | RCT, DB, 5 weeks, industry-sponsored trial (Novartis) | 20mg – 30mg – 40mg – placebo | Self-rated symptoms; investigator-rated symptoms; QOL |
| Medori 2005<sup>7</sup> | OROS methylphenidate | RCT, DB, 5 weeks, industry-sponsored trial (Janssen) | 18mg – 36mg – 72mg – placebo | Self-rated symptoms; investigator-rated symptoms; QOL |
| Casas 2008<sup>8</sup> | OROS methylphenidate | RCT, DB, 13 weeks, industry-sponsored trial (Janssen) | 54mg – 72mg – placebo | Self-rated symptoms; investigator-rated symptoms; QOL |
| Huss 2010<sup>9</sup> | Long-acting methylphenidate | RCT, DB, 9 weeks, industry-sponsored trial (Novartis) | 40mg – 60mg – 80mg – placebo | Self-rated symptoms; investigator-rated symptoms; peer-rated symptoms |
| Weiss 2014<sup>5, 10</sup> | Controlled-release methylphenidate | RCT, DB, 4 weeks, industry-sponsored trial (Purdue Pharma) | 25mg – 45mg – 70mg – 100mg – placebo | Self-rated symptoms; investigator-rated symptoms; QOL; peer-rated symptoms |

*DB= double blind, OROS = Osmotic-controlled Release Oral delivery System, RCT = randomised clinical trial, QOL = quality of life.*

We analysed four efficacy outcomes: Self-rated ADHD symptoms (5 trials, 1807 participants) (**Figure 1**), investigator-rated ADHD symptoms (5 trials, 1904 participants) (**Figure 2**), quality of life (4 trials, 1158 participants) (**Figure 3**), and peer-rated ADHD symptoms (2 trial, 879 participants) (**Figure 4**). We did not observe a dose-response relationship for any of the four outcomes and the mean differences between the methylphenidate groups in each trial were small and most likely clinically irrelevant.

**Figure 1.**
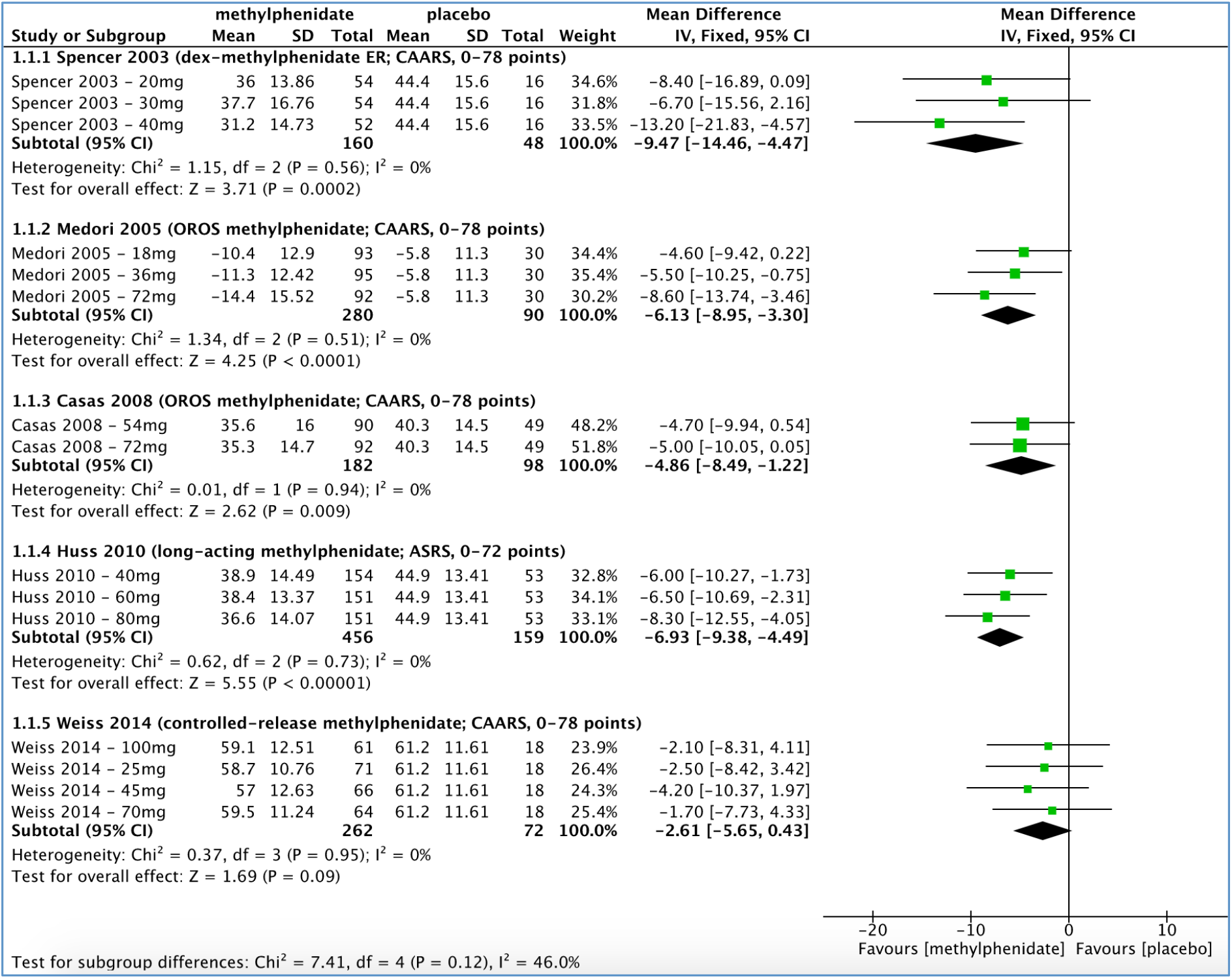
Self-rated ADHD symptoms

**Figure 2.**
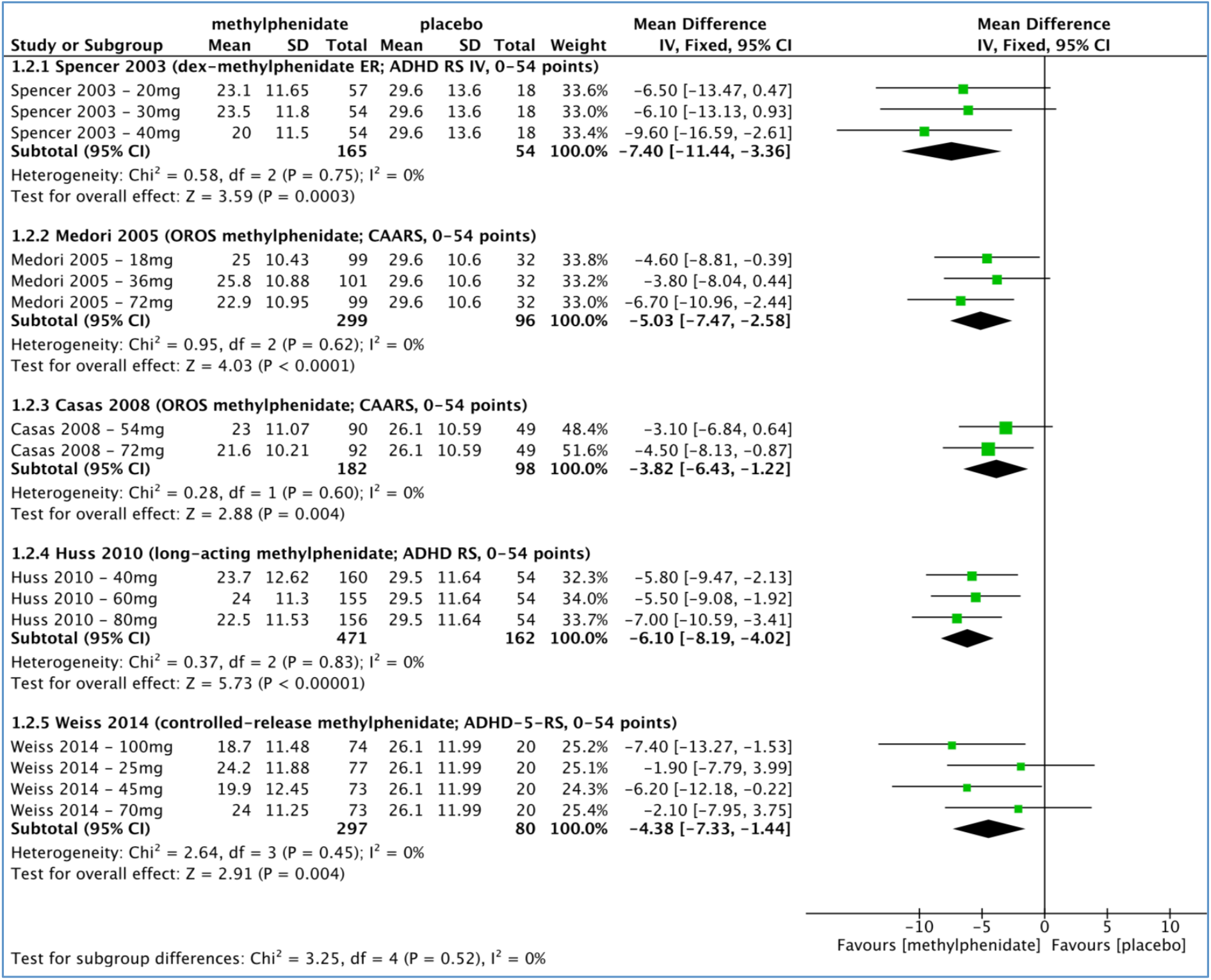
Investigator-rated ADHD symptoms

**Figure 3.**
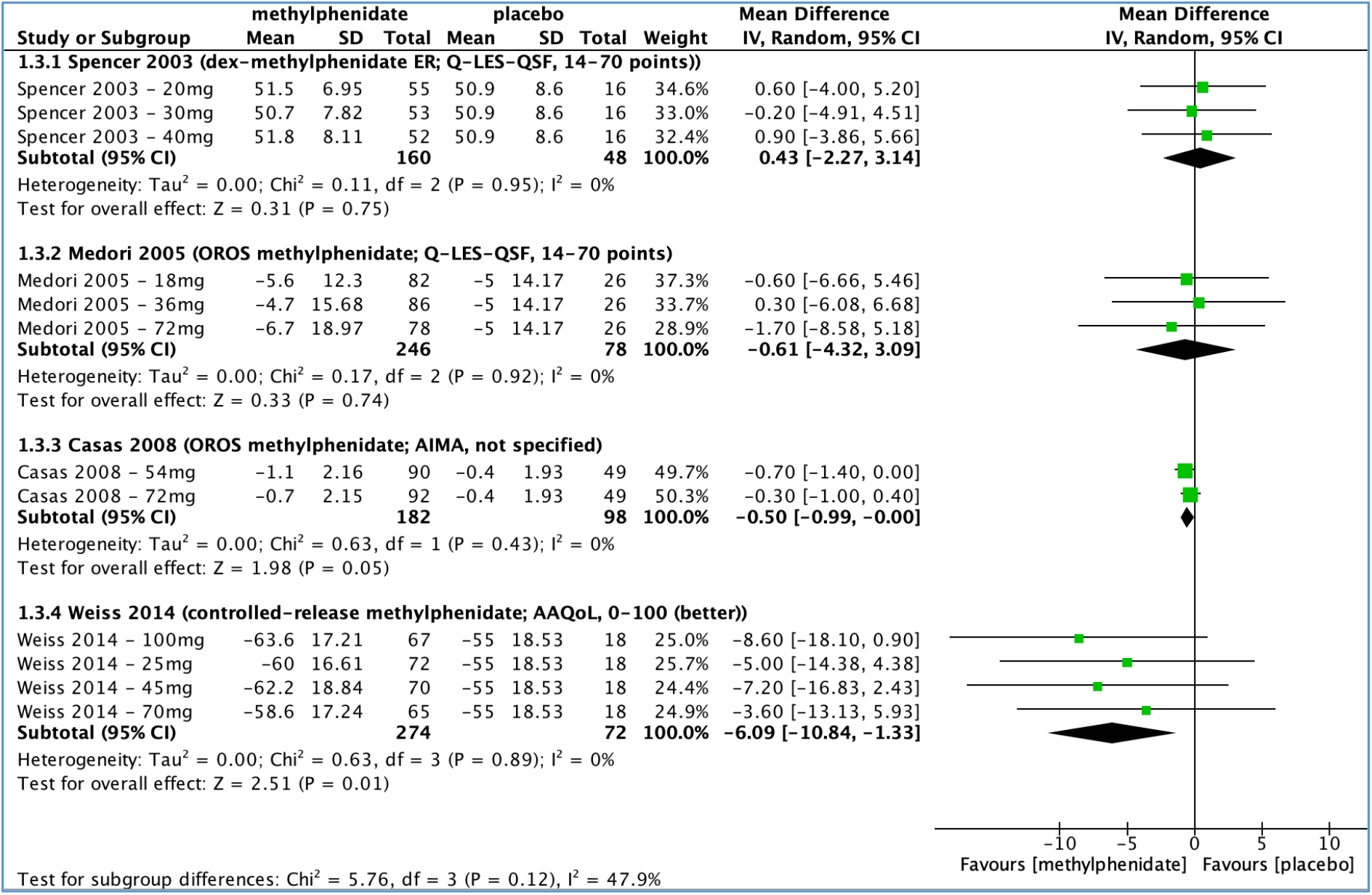
Quality of life

**Figure 4.**
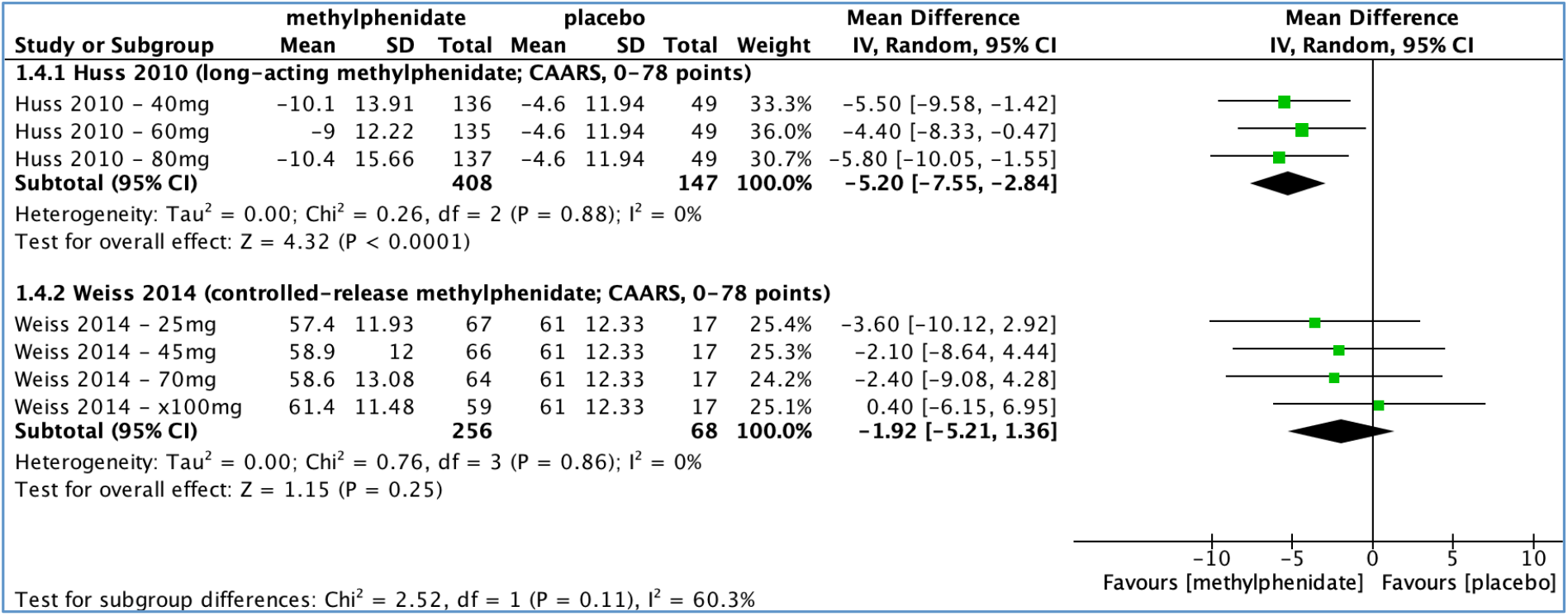
Peer-rated ADHD symptoms

The largest mean difference observed was a 6.5-point difference on a self-rated symptom scale (range 0 to 78 points) in Spencer 2003^6^ between 30 mg and 40 mg dexmethylphenidate. For investigator-rated ADHD symptoms, the point estimates favoured the highest dose in all five trials, but the group differences were only a few points ranging from 1.5 to 5.5 points on a scale from 0 to 54 points. There is limited empirical knowledge on the minimal clinically relevant differences for ADHD rating scales, except for the quality of life outcome scale, Q-LES-QSF, which is estimated to be 3 points.^11^

## Discussion

To our knowledge, this is the first systematic assessment of dose-response relationships of methylphenidate for ADHD in adults. Our analysis was not preregistered, which is a limitation, but data were systematically collected for our review.^4^ It is uncertain whether the trials were powered to demonstrate group differences but the mean differences between the dose groups were small lending no support to dose-response relationships. Other methods may be used to address this question, such as meta-regression, but it was not feasible due to the use of different methylphenidate formulations and doses in this analysis.^12^ Notably, one method should not be used: a comparison across dose titration studies using the mean or median dose, which is unreliable because of the ecological fallacy.^13^ A systematic review^14^ of amphetamines for adult ADHD also did not find differences between ‘lower’ or ‘higher’ doses amphetamines versus placebo, although the used methodology was unclear.

We did not analyse harms due to potential selective reporting. As seen in our dataset, only a few harms outcomes were reported separately for the methylphenidate groups. In a comment^15^ on the most recently approved methylphenidate formulation, we highlighted that the FDA did not authorise the highest dose (100 mg controlled-release) as they found a “disproportionate” increase of certain adverse events compared to the lower doses (25, 45 and 70 mg).

In the letter,^1^ the author further wrote, “the presumed optimal dose” is around 1 mg/kg. The supporting narrative review^16^ stated, “Doses around 1 mg/kg of methylphenidate are correlated with better efficacy, yet are rarely achieved in studies of adult patients”. The statement seems illogical; if you can’t “achieve” a given dose, i.e. titrate participants to a higher dose - likely because of adverse effects – it will obviously not be better than a lower dose. The narrative review (published in 2018) cited two methylphenidate systematic reviews; one that had been retracted in 2016 because of methodological shortcomings,^17^ making it an unfortunate choice of reference; and the other review^18^ used the inappropriate method of comparing mean doses across dose titration trials.

Finally, the assessed pivotal trials have serious methodological limitations, which would likely favour dose-response relationships. Most trials selected their participants based on their response to previous central stimulant exposure,^4^ and selective outcome reporting likely also favours a dose-response relationship, as it would be in the companies’ interest to demonstrate such an effect. Inappropriate handling of missing data such as using the last observation carried forward (LOCF) method could also drive differences between the groups, as dropout rates and reasons for dropouts would vary, and as more dropouts are expected with higher doses. In two of the included fixed-dose trials, Casas 2008^8^ and Huss 2010,^9^ there was likely such inappropriate handling of missing data: For Casas 2008’s highest dose (72 mg) and Huss 2010’s two highest doses (60 mg and 80 mg), there were more participants designated as “responders” than actually completed the trials. The British drug regulator also made this observation about Casas 2008 in their public assessment report (p. 45).^19^

In conclusion, our results do not support a dose-response relationship of extended-release methylphenidate for adult ADHD.

## Data Availability

The full dataset: https://osf.io/wrf65/?view_only=ef3340ea339c44bdad5f83732cf4ba84.

## Acknowledgements

None

## Data sharing statement

The full dataset: https://osf.io/wrf65/?view_only=ef3340ea339c44bdad5f83732cf4ba84.

